# The experiences, challenges, and enablers for promoting interprofessional education among medical students: A Scoping Review

**DOI:** 10.1101/2025.08.13.25333643

**Authors:** Maxwell Ateni Assibi, Bruce Abugri Ayabilla, Patience Afua Adwaapa Karikari, Shamsu-Deen Ziblim, Victor Mogre

**Affiliations:** Department of Health Professions Education and Innovative learning, University for Development Studies.; Department of Population and reproductive health, school of public health, University for Development Studies.

**Keywords:** Interprofessional education, medical students, experiences, challenges, enablers, teamwork, healthcare collaboration

## Abstract

**Objective:** This scoping review maps the literature on the experiences, challenges, and enablers of interprofessional education among medical students.

**Methods:** A scoping review methodology based on the Arksey and O’Malley framework of 2005 was conducted. The search focused on peer-reviewed articles published in English between 2014 and 2024 of various designs (Qualitative, Quantitative and mixed methods). A systematic search of the literature was made through databases like PubMed, Scopus, Web of Science, and Google Scholar using terms like “Interprofessional Education,” “medical students,” “experiences,” “challenges,” and “enablers.”

**Results:** After reviewing 1,500 articles, 11 studies were included, revealing that interprofessional education enhances teamwork and communication through simulation, clinical placements, and peer learning. However, challenges such as scheduling conflicts, resource limitations, and faculty unpreparedness hinder implementation. Effective enablers, including faculty training, structured feedback, and institutional commitment, support engagement and sustainability.

**Conclusion:** Interprofessional education is a crucial tool for preparing medical students to practice collaboratively in healthcare. With the adoption of such established and sustainable IPE programs, the future health care providers will be well equipped to deliver quality, collaborative patient care. Further studies are needed to investigate long-term outcomes of IPE and how it impacts patient care across various settings of health care.

## Introduction

With the increasing complexity of healthcare delivery, interprofessional education (IPE) has emerged as a key strategy for the education of healthcare professionals. The World Health Organization (WHO, 2010) has defined IPE as, “The process by which students from two or more professions learn about, from and with each other to enable effective collaboration and improve health outcomes. Interprofessional education improves collaboration in health care provision towards improved patient care, organization, and health outcomes (Barr, 2013). Early introduction to IPE among medical students is important as the students will be the ones carrying out clinical practice in patient care (Frenk et al., 2010). Interprofessional competencies are also needed to be acquired while pursuing medical training because this will equip the students with the ability to work in interprofessional teams to improve health care delivery (Thistle Thwaite, 2012).

While the advantages of IPE have been widely recognized, its inclusion in medical education remains variable. Its wide-scale implementation is limited by several structural and institutional factors, including professional hierarchies, role ambiguity, and logistical limitations related to faculty preparedness and scheduling (Ahmady et al., 2020; Berger-Estilita et al., 2020; Thompson et al., 2020). Existing traditional discipline-specific curricula further adds to the fragmentation of interprofessional learning, highlighting the necessity of a more cohesive educational approach.

Given their crucial role in healthcare delivery, this review focuses on medical students. As future physicians, they are expected make central clinical decisions and patient care. As future leaders and educators, the need for medical students to be able to work effectively within interprofessional teams becomes paramount due to the need to reduce medical errors and improve patient outcomes through early exposure to collaborative practice. With healthcare systems continuing to move toward integrated, team-based care, training medical students in interprofessional collaboration skills is a matter of great concern.

This literature on IPE has many gaps, as there is limited exploration of medical students’ lived experiences, including the challenges and enablers of interprofessional learning contexts (Bogossian et al., 2023). The majority of the studies are faculty- or institution-focused, which restricts the understanding of students’ perceptions and engagement. Moreover, few studies have evaluated the long-term effect of IPE on students’ career readiness and patient care. These gaps must be addressed to inform strategies for improving IPE implementation and effectiveness.

This scoping review aims to provide a synthesis of the existing evidence pertaining to medical students’ participation in IPE programs, detailing the challenges and enablers to their inclusion. This review will analyzes relevant empirical studies to identify evidence based factors specifically for the implementation of IPE in a range of educational and clinical settings. It will also explore how these differences in IPE exposure are mediated at an institutional level by how institutional structures, professional hierarchies and resource availability affect IPE exposure to inform future interprofessional education.

### Objective of the review

To map existing literature on the experiences, challenges, enablers of medical students in interprofessional education (IPE) programs.

### Research Questions

1. What are the experiences of medical students in interprofessional education (IPE) programs?
2. What challenges do medical students face in interprofessional education programs?
3. What factors promote effective IPE programs?

## Method

A very strong methodological approach was used in the scoping review, which highlighted the results’ credibility, dependability, and reliability. With the help of Levac, Colquhoun, and O’Brien’s (2010) suggestions and the methodology developed by Arksey and O’Malley in 2005, the review aims to conduct a comprehensive mapping of the literature on IPE for medical students. In order to improve repeatability and expand our knowledge of student experiences, challenges, and support in pursuing IPE studies, this technique offers a chance to increase transparency. The PRISMA extension for Scoping Reviews (PRISMA-ScR) by Tricco et al. (2018) serves as the foundation for comprehensive reporting, which includes 20 reported elements and two optional items.

### Study design

In this case, a scoping review approach was used because it is best suited for broad research issues, can support a variety of study designs, and can identify gaps in the literature. Future research and practice may benefit from the framework it provides for synthesizing findings from qualitative, quantitative, and mixed-methods studies (Arksey & O’Malley 2005; Levac, Colquhoun, O’Brien 2010; Tricco et al. 2018). According to Arksey and O’Malley (2005), scoping reviews are particularly suitable for mapping the quantity, scope, and kind of research effort in the field, finding evidence gaps, and elucidating important ideas. This approach is flexible and inclusive allowing the inclusion of wide ranging study designs and methodologies, necessary for tackling complex and various research questions (Levac et al., 2010; Tricco et al., 2018).

### Eligibility criteria

To ensure the research objectives always tally and focus on the relevant literature, some selection criteria were chosen. This is considered as some of the eligibility criteria:

### Inclusion criteria

Peer-reviewed articles from 2014 onwards to 2024.Studies exploring medical students’ experiences, challenges and enablers in IPE settings. Original articles reporting original data using qualitative, quantitative, or mixed-methods designs. Studies conducted in resource-limited or diverse educational settings. Publications available in English.

### Exclusion criteria

Studies focusing solely on healthcare professionals or non-medical students. Commentary, opinion pieces, editorials, or conference abstracts. Articles not reporting empirical data or lacking relevance to IPE among medical students.

### Information sources and search strategy

A multi-database search approach was applied using databases such as PubMed, Scopus, Web of Science, and Google Scholar. The keywords used were “Interprofessional Education,” “medical students,” “experiences,” “challenges,” and “enablers.” Boolean operators (AND, OR) and MeSH terms were used to make the searches comprehensive. The reference lists of the included articles were scanned manually to complement the findings from the database searches.

### Study selection

After one reviewer, MAA, eliminated duplicates from the search results and integrated them in EndNote 7, 2013, another, BAA, verified the procedure. Titles and abstracts were checked independently by two reviewers PAAK, for works concentrating on the design, implementation, assessment, or evaluation of IPE involving medical students. Primary studies from a variety of situations were included in the research to ensure their relevance to the goals of the investigation. The screening findings were compared, and any discrepancies were discussed and settled. After initial screening, full texts of possibly acceptable publications were acquired to allow for a more thorough evaluation. The team members, MAA-PAAK, examined the whole texts to make sure they met the inclusion requirements and that at least one aspect of IPE such as design, implementation, assessment, or evaluation was included. BAA served as an arbitrator to settle any disputes or ambiguities between the two team members. Only those papers having primary data advanced to the data charting step of this evaluation. These suitable articles were then divided into thematic themes based on experiences, difficulties, and facilitators. As a result, careful selection was made to guarantee that 11 excellent publications offer insightful information about the goals of the study.

### Charting the data

For this stage, three reviewers (MAA, PAAK, and BAA) systematically re-reviewed each paper selected for inclusion, confirmed relevance, and charted implementation data based on pre-determined components. Where possible, data not explicitly reported in the papers were extrapolated. Each paper was subsequently reviewed again by BAA and PAAK to ensure all components relevant to IPE implementation were captured.

Trends, insights, together with lessons and challenges from studies’ authors were charted. It provided an organized process where every finding may become relevant within broader understandings of experiences, challenges, and enablers regarding IPE among medical students.

### Collating, summarizing and reporting the results

To guarantee thorough synthesis, data mapping and summarization from the included research were completed using an interactive, iterative process. The three main goals of the research study experiences, obstacles, and facilitators of IPE among medical students, formed the framework for the analysis. In order to find patterns and themes, this was guided by thematic analysis frameworks developed by Stemler (2001) and Tashakkori & Teddlie (2010), who used the education components proposed in the IPECPCP framework by D’Amour & Oandasan (2005).

## Results

A systematic search through databases yielded an initial 1,500 records. The study selection process is shown in figure **1**. Duplicates of 1,440 were eliminated through EndNote software and manual screening as well. A total of 60 unique records were revealed to be worked on. Screening of title and abstracts eliminated 48 records.

**Figure I.**
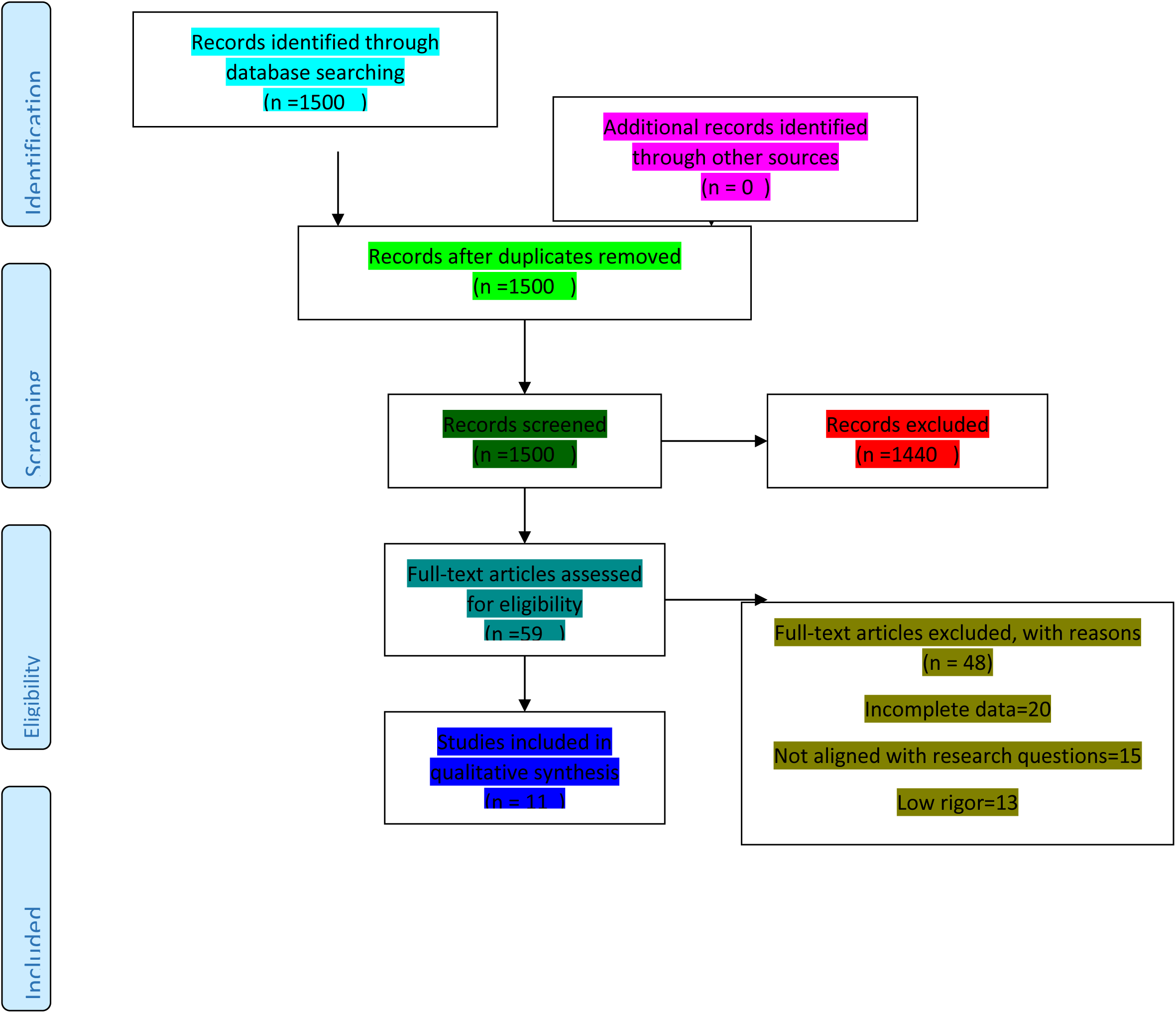
shows the flowchart of the study selection protocol.

There was additional exclusion of 20 more records of incomplete data or the absence of empirical evidence. Another 15 records were also excluded since they were only non-medical students. There was additional removal of 13 records due to low quality or non-applicability to IPE among medical students.

Afterwards, 12 full-text studies were screened against eligibility. One article was further excluded after reviewing full text due to not examining IPE outcomes among student non-students or presenting insufficient background regarding medical students’ experience, barriers, and facilitators.

Eleven studies passed the final synthesis for inclusion as per meeting inclusion criteria. Different geographical contexts as well as diversified study designs involving qualitative, quantitative, and mixed-method designs represented the studies included.

### General characteristics of included articles

The 11 studies that were analyzed had variation in sample size, geographical location, and study design. Table **1** presents the general characteristics of the included articles. Sample sizes ranged from 10 to 683 participants from different countries, including Norway (Reime et al., 2022; 262 participants), Spain (González Blum et al., 2022; 135 participants), Switzerland (Berger-Estilita et al., 2020; 683 participants), Pakistan (Sana Jabbar et al., 2023; 247 participants), Iran (Ahmady et al., 2020; 15 participants), the United Kingdom (Samuriwo et al., 2020; 12 participants; Thompson et al., 2020; 300 participants), Finland (Kangas et al., 2021; 30 participants), China (Xing et al., 2024; 10 participants), Indonesia (Ernawati & Utami, 2020; 588 participants), and the United States (Rivera et al., 2019; 14 participants).

**Table 1.**
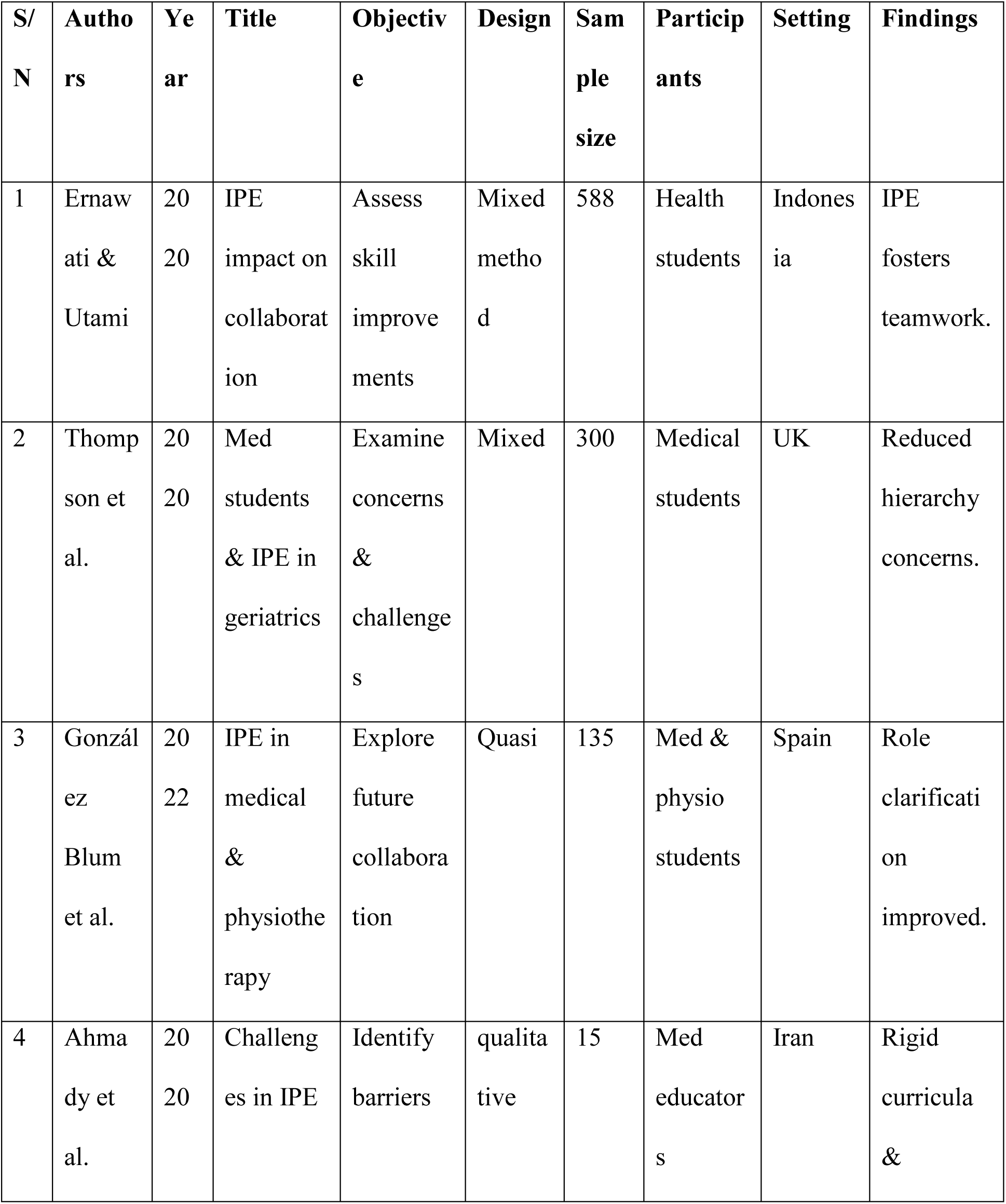
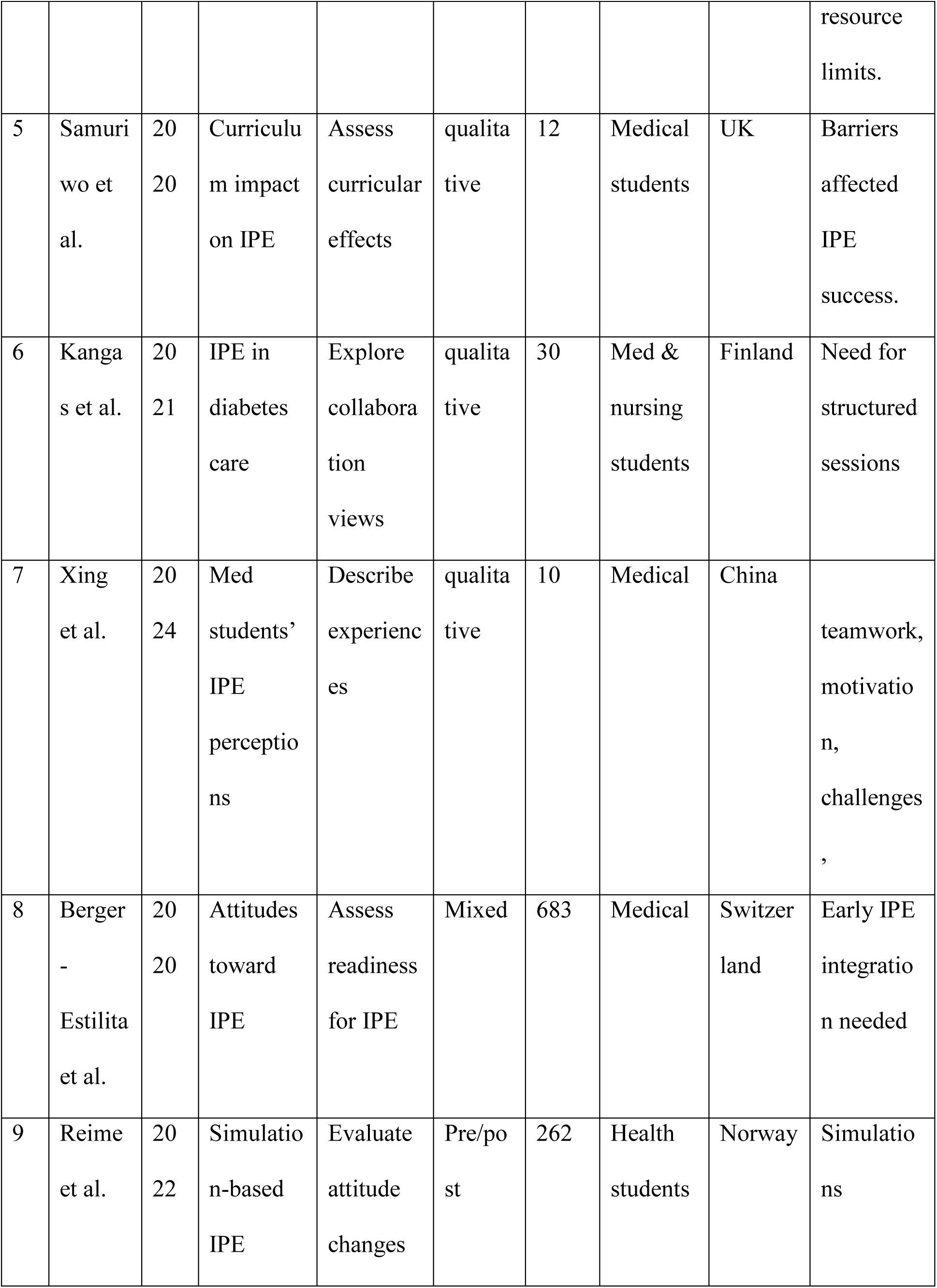
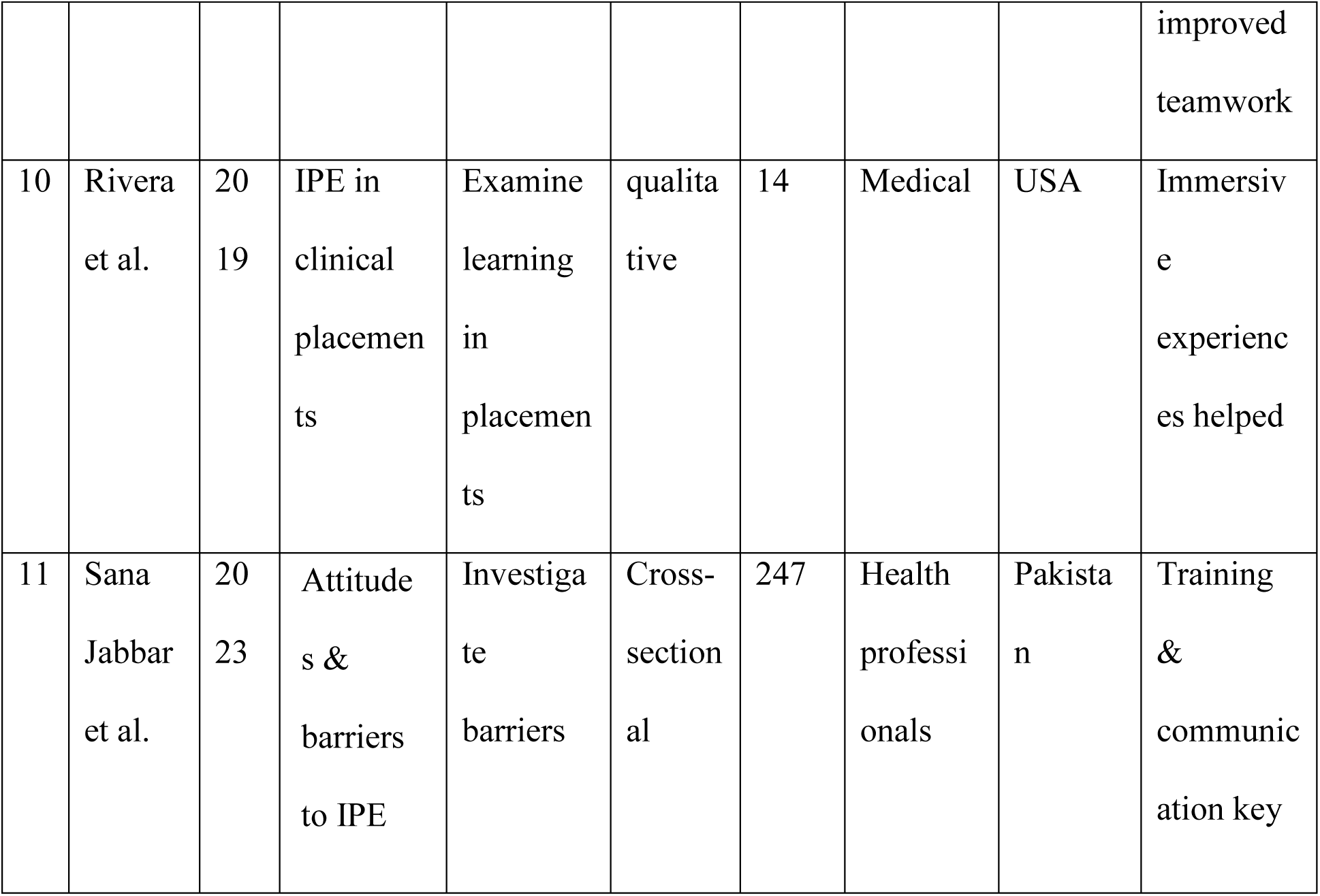
General characteristics of the included studies.

The study design included pre/post-test studies (Reime et al., 2022; González Blum et al., 2022) and cross-sectional surveys (Berger-Estilita et al., 2020; Sana Jabbar et al., 2023). In addition, four studies utilized qualitative approaches including interviews and focus groups that were employed to measure students’ views and issues for IPE (Ahmady et al., 2020; Samuriwo et al., 2020; Kangas et al., 2021; Xing et al., 2024). Three used mixed-method designs whereby more than two strategies in specific, viz. surveys, focus groups, and clinical placements were utilized for examining more than one outcome of IPE (Ernawati & Utami, 2020; Thompson, River et.2019).

### Forms and components of IPE implementation in included studies

Throughout all these studies chosen here, there were diverse structured components and strategies used to develop teamwork among health students in implementing IPE. Most frequent use of this type was simulation training, where students mimicked clinical cases within an artificial setting to allow for the facilitation of teamwork and communication development skills (Berger-Estilita et al., 2020; Reime et al., 2022). There were also clinical placements simulated wherein the students went on interprofessional ward rounds and practicing patient care together with other practitioner professionals (Rivera et al., 2019).

Systematic role-play, active debate, and simulation case management were employed to familiarize students with various professional roles and also collaborative decision-making (González Blum et al., 2022). Small group discussion, peer teaching, and group-based problem-solving exercises were prioritized highly through means of various studies in maintaining interprofessional learning in more relaxed settings (Xing et al., 2024; Berger-Estilita et al., 2020).

Institutional support and staff preparation were key determinants that facilitated effective IPE. These included preparing teachers to conduct IPE sessions and developing specialized interprofessional learning space and curriculum integration for interprofessional experiences (Samuriwo et al., 2020; Ernawati & Utami, 2020). All these implementations demonstrate the diverse but specific measures taken in each study to integrate IPE effectively in health professions education.

## Key findings

The review identified three core themes relating to medical students’ experiences, challenges and enablers of interprofessional education (IPE). The findings of the themes are summarized in table 2.

**Table 2:**
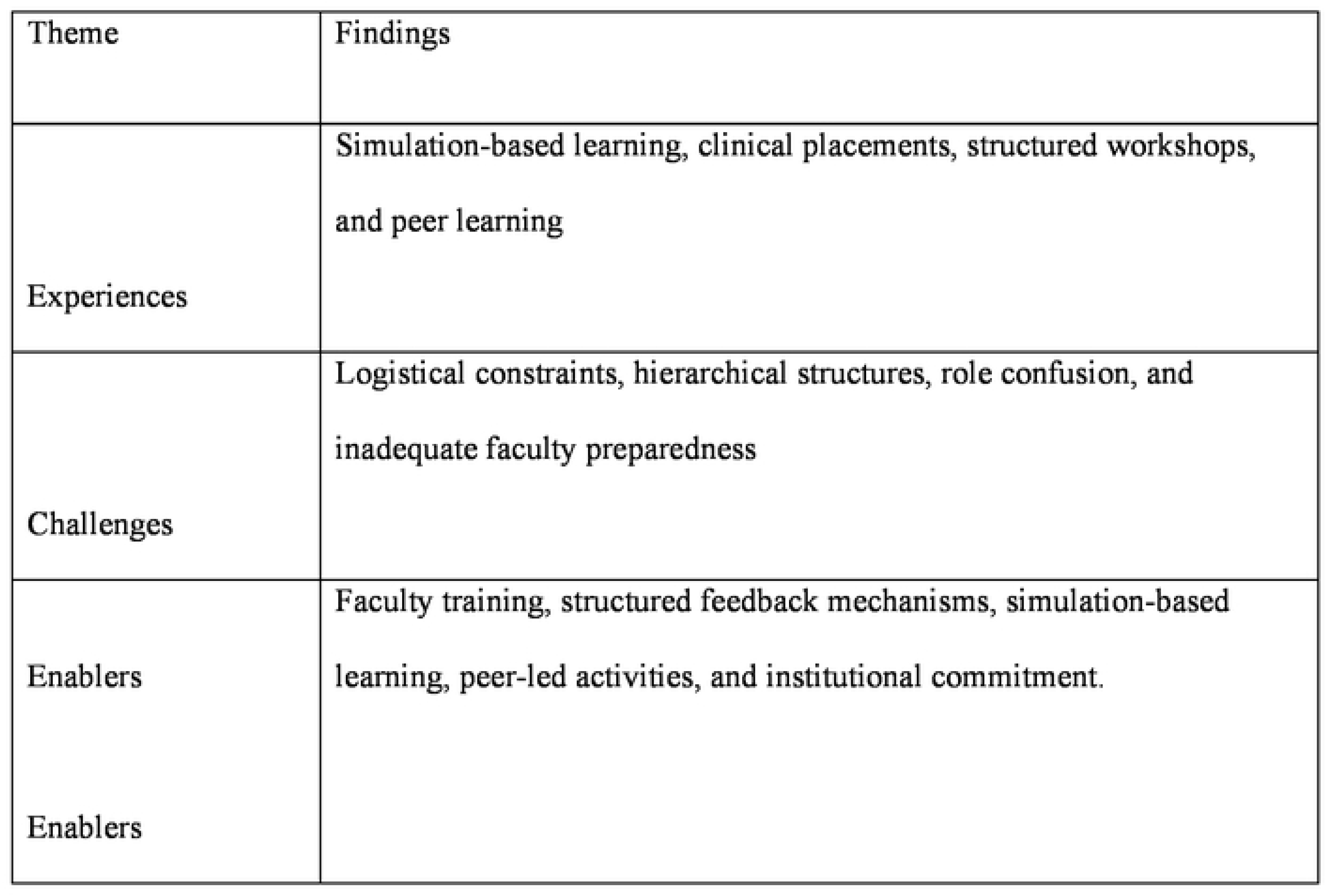
Summary of findings.

### Experiences

Five studies focused on the medical student’s experiences during IPE (Reime et al., 2022; Rivera et al., 2019; González Blum et al., 2022; Berger-Estilita et al., 2020; Thompson et al., 2020). According to the evidence from the review, simulation-based education aided students to face actual clinical aspects of the real world, providing opportunities to improve decision making in case of emergencies. Reime et al. (2022) also showed that the students exposed to simulation training were more competent in managing emergencies and more contiguous with patients. Berger-Estilita et al. (2020) found that students appreciated the ability to identify, in a simulated environment, processes they would later encounter in real clinical practice.

Clinical placements were one of the experiences that were completely characterized as life changing for medical students. Rivera et al. (2019), González Blum et al. (2022) created the extent to which involvement in multidisciplinary ward rounds and patient consultations enhanced students’ experiences of interprofessional patient care. For most students, their experience of working with pharmacists, nurses, and physiotherapists was at the Centre of them knowing the other perspectives and skills that were essential to care for patients.

The review also established that structured workshops had a very central position in the medical students’ experiences in terms of IPE. In accordance with González Blum et al (2022), integrating role-play exercises of mock case discussions in workshops exposed the students to unique healthcare team experiences. Structured events allowed students to feel confident in clinical decision-making and capable when interacting with other health providers.

The second fundamental component of IPE experiences among medical students from the studies was peer learning activities. Berger-Estilita et al. (2020) found that students who participated in peer-assisted group discussions and case-based learning, developed a better understanding of solving problems globally in interprofessional teams. A few students mentioned that learning from peers from other disciplines allowed them to fill knowledge gaps and became more effective collaborators in future clinical practice.

Though these are positive experiences, the review also found some medical students struggled to settle into interprofessional practice. Thompson et al. (2020) and Jabbar et al. (2023) highlighted that professional boundary dilemmas emerged when students were first faced with interdisciplinary settings.

### Challenges

Five of the reviewed studies (Ahmady et al., 2020; Jabbar et al., 2023; Reime et al., 2022; Kangas et al., 2021; Berger-Estilita et al., 2020) recognized the challenges faced by medical students in the implementation of IPE programs. Out of which, two studies pointed out logistical challenges specifically around scheduling and limited resources. Jabbar et al. (2023) and Ahmady et al. (2020) showed that the highly demanding schedules in medical education made coordination of learning activities across the disciplines challenging for medical students due to suboptimal timetables and a lack of institutional support.

The review also established hierarchical structures in medical learning environment being a formidable challenge to engaging in effective IPE programs. Based on Reime et al. (2022) and Kangas et al. (2021) findings, some students perceived themselves in the highest level of the healthcare team and thus had unbalanced power bases that did not promote quality partnerships with other healthcare professionals. Role confusion was also part of the issue medical students protested based on the review, with Thompson et al. (2020) having identified students’ confusion since they performed diverse roles in interprofessional teams. This led to role confusion, which mainly caused uncertainty and disinterest. Finally, inadequate preparation by faculty was considered a challenge, with Berger-Estilita et al. (2020) and Rivera et al. (2019), indicating that, the faculty did not have enough skills and experience to use interprofessional learning appropriately and this affected IPE programs.

### Enablers

Seven most influential enablers of effective IPE implementation were determined in seven studies (Berger-Estilita et al., 2020; Ernawati & Utami, 2020; Jabbar et al., 2023; Reime et al., 2022; Rivera et al., 2019; Samuriwo et al., 2020; Xing et al., 2024).

Support and faculty development was a prime facilitator of IPE implementation based on the review. Ernawati & Utami (2020) ascertained systematic faculty development programs promoted facilitators to effectively facilitate interprofessional learning opportunities. One such key facilitator was instituting systematic mechanisms of feedback. That was purportedly proven by Jabbar et al. Jump (2023) based on the use of feedback by faculty, where students who received periodic feedback had significant improvements in confidence and collaboration skills. As Reime et al., (2022), concluded, simulation-based learning activity was recognized as a facilitator of IPE. They also indicated high-fidelity simulation improved students’ interdisciplinary team-working ability significantly. Student-led case discussion and group project were also found to work, as was Xing et al. (2024), which determined how peer relationship helped facilitate respect and professional development. Samuriwo et al. similarly indicated institutional commitment as a significant facilitator, too. They identified that institutions with IPE being added to curricula and with committed resource allocation for interprofessional learning space had higher student participation and program viability. Institutions that facilitated success of such IPE programs were those that referred to helped demystify the value of collaborative methods to education in medicine to help gain support for long-term IPE programs.

## Discussion

The review went further to emphasize that interprofessional education (IPE) provides medical students with extensive experiential learning opportunities (Reime et al., 2022; Berger-Estilita et al., 2020). Simulation-based learning was one of the approaches that was used comprehensively to expose the students to clinical cases. Studies conducted by Reime et al. (2022) and Berger-Estilita et al. (2020) note that apart from enhancing the confidence of students to manage patient cases, that simulation-based training strengthens their ability to perform in interprofessional teams. The study notes that students subjected to this type of simulation training demonstrated significantly more mastery of some skills in terms of listening to colleagues, responding to colleagues, and assessing and treating patients in emergent situations. High-fidelity technology simulations gave students the opportunity to develop interprofessional collaborative practice approaches in a simulated environment prior to exposing students to working in real clinical placements thereby facilitating the increasing importance of teamwork to care for patients. Nonetheless, the effectiveness of simulation-based learning was reported to depend on the quality of facilitation and support in institutions, indicating that further studies may need to be done to increase its use in IPE programs (Ernawati & Utami, 2020).

Another experience reported for medical students was clinical placements (Rivera et al., 2019; González Blum et al., 2022). Interprofessional ward rounds and face-to-face patient care interactions provided students with valuable and rare opportunities to learn about other healthcare professionals’ roles and responsibilities. The studies concluded that students who experience collaborative clinical placements understood patient management more holistically and experienced team-based decision-making (Samuriwo et al. 2020). Clinical placement programs were also linked to an enhanced recognition of the value of non-physician professionals including nurses, pharmacists and physiotherapists in patient care, and exposed the way that IPE specifically could enrich interdisciplinary teamwork. However, due to a lesser integration of the disciplines in the healthcare setting, the interprofessional learning in clinical placements was extended. In some studies it was reported that in settings that did not practice interprofessional collaboration, students were not able to engage fully in team-based practices which reinforces structured integration of IPE in clinical settings (Jabbar et al., 2023).

Medical students experiences were also characterized based on workshops and peer-learning activities that were also substantial (González Blum et al., 2022; Berger-Estilita et al., 2020). The students were exposed to their roles on the healthcare team through case-based workshops and role-plays. These interactive learning methodologies utilized promoted the growth of critical analysis, teamwork and collaboration in problem-solving, and enhanced communication with other health care practitioners (Ahmady et al., 2020). By learning from one another it allowed for students with diverse specialties to coach each other and learn from the other’s lenses. In one study, active peers within peer-led IPE sessions wished for greater decision-making and agreement by consensus (Xing et al., 2024). However, heterogeneity of student level of experience and variability in participation of students within the peer learning activities were found, and additional studies must be performed to decide effective strategies with which to implement these pedagogical interventions (Thompson et al., 2020).

The review identified a list of obstacles in the efficient practice of IPE. Problems with logistics, such as clash timetables and resources within institutions not being accessible, were invariably reported throughout various studies (Ahmady et al., 2020; Jabbar et al., 2023). From the findings of the said studies, variations in timetabling by schools for lecturers in the health disciplines tightly limited the student possibilities for involvement in interprofessional learning activities. Poor integrated planning made it difficult for students to engage in working together, producing a limited cumulative impact of IPE (Samuriwo et al., 2020). Other structural impediments included having access to adequate physical and technological infrastructure for carrying out IPE activities, contributing to participative challenges. These findings implied the need for institutions to provide adequate attention to developing flexible models of timetabling, and investing in infrastructure that would facilitate interprofessional learning (Rivera et al., 2019).

Medical students’ training was also perceived to have hierarchical structures that prevented interprofessional education from succeeding (Reime et al., 2022; Kangas et al., 2021). The medical students perceived superiority over other health professionals and hence there was a power imbalance among interprofessional groups. Such belief caused decision-making to be dominated by medical students while other professionals struggled to join in such decision-making (Thompson et al., 2020). The traditional hierarchical model of medical education facilitated such dynamics and asymmetric involvement in cooperative learning activities (Berger-Estilita et al., 2020). This can be done only by changing the culture of health education from cooperation, respect, and interdependence (Ernawati & Utami, 2020).

Role confusion was a significant problem in the review (Thompson et al., 2020). In order for medical students to consider not only their own role within interprofessional teams, but also their own position within this, ideas were sometimes confused and participation became less frequent. This was particularly the case where the students had little or no exposure to interprofessional working prior to their IPE experience (Jabbar et al., 2023). When students were not definite about their roles, they were not self-assured in enabling team-based care, hence limiting chances for interprofessional learning (González Blum et al., 2022). The research emphasizes the significance of engaging formal role clarification activities in IPE programs so that students are definite about their roles and feel empowered to engage in collaborative patient care (Samuriwo et al., 2020).

Institutional and pedagogic features were viewed as necessary to enable IPE. Ernawati & Utami (2020) viewed professional development and training of the faculty as central enablers of the achievement of interprofessional learning. Research also recognized that faculty members specializing could enable successful implementation of interprofessional education. These specially trained faculty exhibited greater success in creating inclusive and interactive learning environments. Institutions that complied with the prescribed faculty training programs reported higher levels of engagement amongst learners, and improved incorporation of IPE content into medical curricula (Jabbar et al., 2023).

Systematic feedback mechanisms were equally paramount in enhancing students learning processes (Reima et al., 2022). Supportive and timely feedback provided students an opportunity to reflect inwardly on their interprofessional skills, to identify the gaps, and to relate learning back to clinical practice in a more realistic manner.

Simulation-based training was a major factor in helping students acquire interprofessional working behaviors in simulation environments prior to them moving to real clinical environments (Samuriwo et al., 2020). High-fidelity simulation helped students practice their verbal and non-verbal communication, leadership, and teamwork skills in an environment that did away with the stress of a clinical setting (Rivera et al., 2019). Xing et al. (2024) implemented peer-directed activities that have been effective in enabling sharing of knowledge and collective decision-making between students from different professional backgrounds. Learning from each other directly helped develop respect for the different professions in health and collaboration in providing patient care.

The review also identified institutional commitment as a major enabling factor for the sustainability of IPE programs. Institutions that integrated interprofessional learning into their curricula and allocated dedicated resources for implementation had higher student engagement and more sustainable programs (Berger-Estilita et al., 2020). IPE infrastructure commitments such as simulation labs, interprofessional training facilities, and collaborative learning areas enable continued student engagement. Samuriwo et al., (2020) and Rivera et al. (2019) pointed out that institutions promoting good IPE policies were assisting in creating more integrated and productive learning experiences, supplementing collaboration in professional healthcare settings.

### Future research directions

Longitudinal studies are required to explore how IPE impacts professional practice or patient outcomes. Even though short-term impacts of IPE (better teamwork, communication, and collaboration) have been studied, longitudinal evidence regarding how these competencies lead to practice and patient care is rare. Longitudinal studies are required to determine the sustainability of IPE outcomes and how they impact improved healthcare delivery and patient safety.

Further research should assess new strategies that consider these gaps and challenges discussed in the review, with a focus on those applicable to resource-poor settings. For example, studies that investigate the benefits that low-cost, simulation-based tools, e-learning modules, and community-based IPE provide may help illustrate resource limitations and infrastructure challenges. A plan for implementing cultural change across academic institutions could also help in reducing professional silos, which in turn could foster stronger collaboration.

Future research should also explore the experiences, challenges and opportunities of health professionals and faculty on the community based education and services as a strategy for promoting interprofessional education.

The role of faculty in IPE is still a very important area of investigation. Future research should explore the best practices for faculty training and development to enhance their competence and confidence in delivering IPE programs. Studies could also examine the impact of faculty mentorship and support systems on the success of IPE initiatives.

Another avenue of importance is the integration of IPE within the general healthcare curriculum. Comparative studies are required to establish how different curricular models, such as early exposure versus later-stage integration and mandatory versus elective IPE programs, fare in comparison. How to smoothly integrate IPE into existing curricula without additional burdens on either students or faculty is critical for its long-term sustainability.

In addition, the future research study needs to reflect on the underrepresentation of some professional health studies. It should therefore be widened to involve other disciplines such as pharmacy, social work, and public health to broaden the scope of Interprofessional collaboration. Such a study that will also take into account the special needs and perspectives of students from diverse cultural and socio-economic backgrounds would help to ensure inclusiveness and equity in IPE programs.

Finally, international research should prioritize studies that include low- and middle-income countries where resource constraints and challenges in healthcare often differ from high-income settings. Such cooperative, multi-country studies might thus provide a wealth of insight into the adaptation of IPE within diverse healthcare systems and populations.

Addressing these research priorities will help move the field of IPE forward while contributing to the development of healthcare professionals prepared to work collaboratively in high-quality, patient-centered care.

### Conclusion

Interprofessional education (IPE) has emerged as a predominant pedagogical strategy to prepare medical students for collaborative practice in healthcare, yet we know surprisingly little about the quality of what is being learned. In this review, we demonstrated that IPE is an effective individual-level learning experience but one that hinges on structured implementation, faculty preparation, and institutional commitment. The other critical areas that need to be anchored include overcoming logistical constraints, empowering faculty members, and developing the collaborative culture required for optimizing IPE outcomes.

Evidence of an effective IPE environment must transcend integration of knowledge and call upon interconnectedness of learning with application in real life, which students need to build its competence within disciplines. Also, institutional policy must promote the development of structured learning experiences with the guarantee that students have adequate resources and mentorship in order to capitalize on IPE programs. Finally, IPE programs will need to be continuously evaluated and modified in order to meet the changing needs of health care.

IPE can lay firm foundations to building the capacity of future health care professionals through targeted implementation efforts in combination with conducive learning environments. If we want to be confident that the health care of the future will be better, we will have to be dedicated to continue development of this education after all, more collaboration in health care can only lead to more good outcomes for our patients.

## Strengths and limitations of the study

Review was of systematic approach, employing the guidelines for scoping review (Arksey & O’Malley, 2005; Levac et al., 2010) and the PRISMA-ScR guidelines to justify methodological rigor and transparency over study selection and synthesis. Geographical heterogeneity is also incorporated in that it has studies from different countries and regions which augments generalizability of evidence across different healthcare settings as well as education settings. Lastly, the review also has a specific targeted analysis of medical students and also a single particularly targeted analysis of their enablers, challenges, experiences in interprofessional education (IPE) as such, it does narrow the literature gap to a certain extent compared to more general health professions’ reviews. However, there limitations: The review utilized only published papers in English language and this potentially excluding relevant findings from other languages. The review had no direct comparisms as a result of the different study designs. Caution should be taken in generalizing the findings of this review since they are constrained by the search conditions.

## Data Availability

N/A

## Acknowledgments

The authors also acknowledge the University for Development Studies, Department of Health Professions Education and Innovative Learning, for the academic support. We also appreciate the provision of the library staff’s work and access to academic databases, which has made this review possible.

